# Urine TIMP2.IGFBP7 reflects kidney injury after moderate volume paracentesis in patients with ascites: A randomized control study

**DOI:** 10.1101/2023.12.20.23300324

**Authors:** Anuchit Suksamai, Sanpolpai Khaoprasert, Amnart Chaiprasert, Sakkarin Chirapongsathorn

**Affiliations:** Division of Gastroenterology and Hepatology, Department of Medicine, Phramongkutklao Hospital and College of Medicine.; Division of Nephrology, Department of Medicine, Phramongkutklao Hospital and College of Medicine.

**Keywords:** decompensated cirrhosis, novel biomarker, urine TIMP2, urine IGFBP7, AKI

## Abstract

**Background:** Patients with cirrhosis undergoing therapeutic paracentesis are at risk of developing kidney injury, but this has not been well evaluated in a non-large volume paracentesis setting.

**Objective:** The aims of this study were to determine the risk and consequence of acute kidney injury (AKI) and its progression in patients with decompensated cirrhosis after moderate volume paracentesis by use of a urine test measuring tissue inhibitor of metalloproteinases-2 (TIMP2) and (IGFBP7).

**Methods:** A randomized, prospective cohort study was performed. All outpatients with decompensated cirrhosis with ascites and diuretic complications were enrolled and randomized into 3 liters and 5 liters of paracentesis groups. Serial urine samples were analyzed for [TIMP2]*[IGFBP7] concentration before and after paracentesis.

**Results:** A total of 90 patients with decompensated cirrhosis were consecutively enrolled during the study period. After screening, 29 patients were enrolled in the 3-liter paracentesis group, and 25 patients were enrolled in the 5-liter paracentesis group. The mean of the MELD score was 8 ± 1.2. Urine TIMP2.IGFBP7>2, rising urine TIMP2, and rising urine TIMP2/urine Cr were shown in patients within the 5-liter group for 48% (p = 0.015), 32% (p = 0.049) and 76% (p =0.010), respectively. There was no statistical difference between the two groups in the rapid decline of GFR, admission, or death.

**Conclusion:** Urine TIMP2.IGFBP7>2 predicted renal tubular injury in patients in the ascites release 5 L group therefore, ascites release 5 L may not be safe. Kidney injury could occur even less than 5 liters of ascites release in decompensated cirrhosis as demonstrated.

The national clinical registration number was TCTR20191116003.

## Introduction

Cirrhosis is a chronic liver disease that is commonly identified in clinical practice. Prevalence of acute kidney injury (AKI) in inpatients with cirrhosis is higher than 20%^1^. Decompensated cirrhosis is likely to increase the risk of renal failure and is associated with decrease in effective arterial blood volume, including diuretic therapy for ascites, abdominal paracentesis, gastrointestinal bleeding, infection or hepatorenal syndrome, in which mortality rate will increase.^1–3^ In patients with liver cirrhosis, discrepancies between serum creatinine levels and renal function can be accentuated by malnutrition, reduced muscle mass, and increased tubular secretion of creatinine. In addition, ascites and peripheral edema can further decrease the creatinine level by widening the distribution of creatinine in the body.^4, 5^ Therefore, creatinine levels that are used in the variables of the MELD score may not be a good predictor of mortality in all patients with cirrhosis.^6^ The discovery of new biomarkers has recently been used in the field of acute kidney injury because of the simple access to urine analysis.^7^ There are some recent studies that have revealed the use of urinary biomarkers such as cystatin C, neutrophil gelatinase-associated lipocalin (NGAL), interleukin 18 (IL-18), kidney injury molecule 1 (KIM-1), and liver type fatty acid-binding protein (L-FABP) for the diagnosis of AKI in cirrhosis.^8–10^

Tissue inhibitor of metalloproteinases-2 (TIMP2) and insulin-like growth factor-binding protein 7 (IGFBP7) are expressed and secreted in primary kidney epithelial cells of proximal and distal tubule origin in cell culture and tissue, where IGFBP7 is secreted mostly from proximal tubule cells and TIMP2 is expressed and secreted mostly by distal tubule cells.^11^ TIMP2 and IGFBP7 are associated with G1 cell cycle arrest during the very early phase of cellular damage such as inflammation, ischemia, oxidative stress, or receiving renal toxicity agents, which can further prevent proliferation of renal tubular cells to the G_2_ and M phases of the cell cycle.^11–13^

According to previous studies, TIMP2 and IGFBP7 are used only in critically ill patients who have been admitted with respiratory illness, cardiovascular disease, neurological disease, surgical problems, septic shock, and trauma and can be used as biomarkers to predict AKI.^14–16^ The result of TIMP2 x IGFBP7/1000 > 2 means that the result is positive. This means that patients have a high risk of sudden renal failure after 12 hours of follow-up (14). However, there is no data on these two biomarkers in patients with cirrhosis.

Large volume paracentesis is an effective treatment for refractory ascites. There is one study suggests that a single 5-liter paracentesis in patients with cirrhosis and tense, diuretic-resistant ascites without albumin infusion is safe and causes no disturbances in systemic and renal hemodynamics 48 hours after the procedure.^17^ However, there have been a small number of patients and no data on early detection of acute kidney injury (AKI). The aim of the study is to evaluate if urinary biomarkers can predict AKI, a reduction in GFR, or changes in MAP after moderate volume paracentesis.

## Methods

### Subjects

A randomized, prospective trial was conducted from December 2018 to December 2021. Eligible individuals were outpatients with cirrhosis aged between 18 and 80 years who were diagnosed with criteria of diuretic-resistance or diuretic-intractable ascites according to a combination of clinical, biochemical, and imaging studies such as ultrasonography, computed tomography, and magnetic resonance imaging, as well as the presence of varices, ascites, or liver biopsy.

All consecutive patients were screened and approached for enrollment by investigators at the Gastroenterology and Hepatology outpatient clinic, Phramongkutklao Hospital, Bangkok, Thailand. Patients must provide written, informed consent before any study procedures occur. Exclusion criteria were patients with end-stage renal disease receiving renal replacement therapy, patients with acute kidney injury during enrollment, patients with unstable hemodynamics during previous abdominal paracentesis, patients with cardiac arrhythmia or heart failure, patients with pacemaker devices owing to the inappropriate measurement of heart rate and blood pressure, shock during enrollment, patients with received radio-contrast media or received nonsteroidal anti-inflammatory drugs within 2 weeks, urinary tract infection, and pregnancy. Patients with deviation from the intended liters of ascites release as randomization, for instance, randomized to 5 liters and only 3 liters came out, were also excluded. All enrolled cirrhotic patients with ascites did not receive an albumin infusion or other volume expander during abdominal paracentesis because no more than 5 liters of ascites were removed. Information on medical history, current use of medications, and diuretic dosage, with the cause of cirrhosis was recorded from each patient. Body weight, height, and calculated body mass index (BMI) as body weight (kg) divided by height (m) squared (kg/m^2^) were measured. Laboratory evaluation includes hemoglobin, platelet count, prothrombin time, and international normalized ratio (PT/INR), albumin, bilirubin, aspartate aminotransferase (AST), alanine aminotransferase (ALT), sodium, blood urea nitrogen and creatinine were collected from all participants. Child-Pugh score (CTP), the model for end stage liver disease (MELD and MELD-Na) scores, and glomerular filtration rate (GFR) according to the chronic kidney disease epidemiology collaboration (CKD-EPI) were performed to assess the severity of liver and kidney impairment. Flowchart of the study was shown in Figure 1.

**Figure 1.**
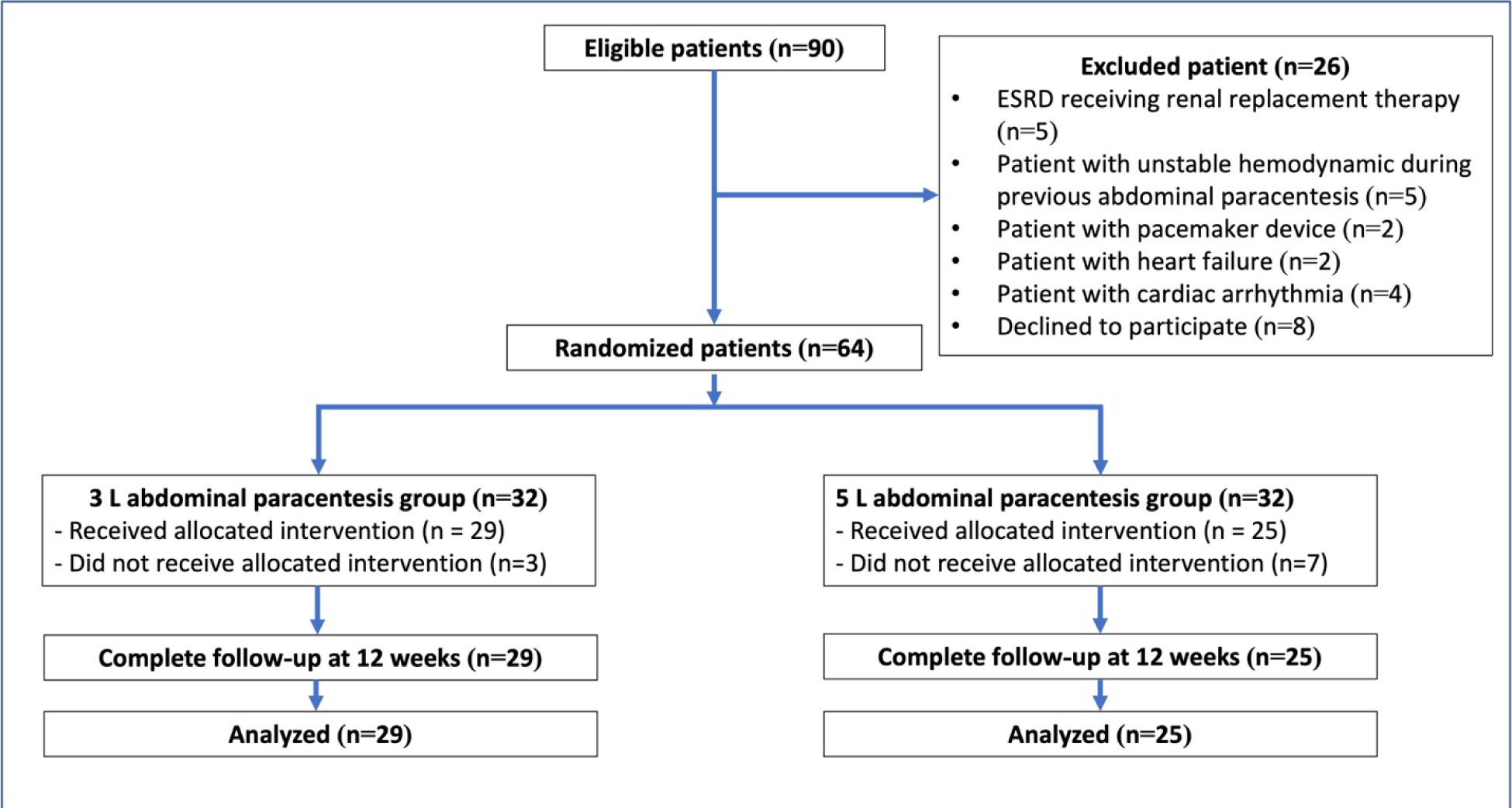
Flowchart of the study

### Study design, randomization, and interventions

The eligible patients were randomized with the use of a computer-generated block of four randomizations and stratified into two groups, which received abdominal paracentesis for 3 liters or 5 liters of ascites. The research assistant generated the random allocation sequence and assigned the participants to two abdominal paracentesis groups. The main investigator, medical laboratory technologist, and patients were blinded to abdominal paracentesis data. Urine samples were collected before and after completing abdominal paracentesis to evaluate the primary endpoint. Blood pressure and heart rate were recorded every 30 minutes until the end of the procedure. Eligible patients were then followed every 2 weeks until 12 weeks, and total abdominal paracentesis times, admission, and death due to cirrhotic complications were recorded to assess the secondary endpoints. Serum creatinine at 2 weeks of follow up was collected to define the acute kidney injury. Serum creatine levels at 12 weeks of follow up were evaluated, and GFRs were estimated by the CKD-EPI equation (ml/min/1.73m^2^) to define the rapid progression of kidney function within 3 months.

This study was approved by the institutional review board and was registered with the national clinical registry; the registration number was TCTR20191116003. All patients provided complete informed consent to participate in the study.

### Sample collection and method of measurement

Urine samples from each enrolled patient were collected before and after abdominal paracentesis and stored at −20°C after being centrifuged at 2,000 x g for 10 minutes. There was no freeze and thaw cycle. No proteinase inhibitors were added to the samples. The samples were examined by a qualified medical technologist. The medical laboratory technologist was blinded to clinical data.

Urine tissue inhibitor of metalloproteinase-2 (TIMP2) and insulin-like growth factor-binding protein 7 (IGFBP7) were detected by sandwich enzyme-linked immunosorbent assay (sandwich ELISA). The sandwich ELISAs quantified antigens between two layers of antibodies (such as capture and detection antibodies). The antigen to be measured must contain at least two antigenic epitopes capable of binding to antibody. The procedure for a sandwich ELISA first required the well of an ELISA plate to be coated with a capture antibody, then the analyte was added, followed by a detection antibody that was conjugated with an enzyme-reagent. A substrate solution was then added to the wells, and color developed in proportion to the amount of antigen-antibody complex. The color development was stopped, and the intensity of the color was measured.

Quantikine^®^ ELISA by Abcam^®^, Cambridge, U.S.A. was used for the detection of urine TIMP2. In this assay, urine as a sample was prepared by combining the sample and calibrator diluent buffered protein in a 2-fold dilution. A monoclonal antibody specific for human TIMP2 had been pre-coated onto a microplate. Then, assay diluent buffered protein base was added to each well. Standards and samples were pipetted into the wells and incubated for 2 hours at room temperature on a horizontal orbital shaker set at 500 rpm. Each well was cleaned and aspirated four times. Conjugated urine TIMP2 was added to each well and incubated for 2 hours at room temperature on the shaker, then repeated the aspiration and wash cycle four times. The substrate solution was added to each well, protected from light, and incubated for 20 minutes at room temperature on the benchtop. Stop solution was added to each well. The optical density of each well was determined within 30 minutes using a microplate reader set to 450 nm. For determining the sample assay, a standard curve was generated with seven standard concentrations. Detailed steps as in the package insert were followed.

SimpleStep ELISA^®^ Kit by Bio-Techne^®^, R&D Systems Inc., Minneapolis U.S.A. was used for detection of urine IGFBP7. In this assay, urine as a sample was prepared by diluting 1:800 in diluent, which was provided in the kit. An anti-tag antibody was pre-coated onto a microplate. Samples and standards were added to each. Then, the mixture of tag-labeled capture antibody and reporter conjugated detector antibody was added to each well and incubated for 1 hour at room temperature on a plate shaker set to 400 rpm. Each well was aspirated and washed three times. A substrate was then added to each well and staked on a plate shaker for 1 minute to mix. The reaction was then stopped by the addition of a stop solution, completing the change of color. The optical density of each well was determined using a microplate reader set to 450 nm. For determining the sample assay, a standard curve was generated with 8 concentrations. Detailed steps as in the package inserted were followed.

The urinary TIMP2 and IGFBP7 were measured in ng/mL and multiplied with each other for increased power of detection. Urine TIMP2 and IGFBP7 were indexed to urine creatinine to adjust for variability in urine concentration. Rising urinary TIMP2 and IGFBP7 levels represent that there was more tubular secretion of biomarkers compared before and after abdominal paracentesis.

## Outcome measurement

### Primary outcome

The primary endpoint was the development of any stage of AKI according to the KDIGO guidelines within 2 weeks of follow up. The reference values for serum creatinine were obtained at the time of enrollment.

**Secondary outcomes** were defined as the following:

- The hemodynamic event was defined by decreased systolic blood pressure ≥ 10 mmHg decreased mean arterial blood pressure ≥ 10 mmHg or a risen heart rate ≥ 20 beats per minute observed within 120 minutes after abdominal paracentesis.
- Rapid decline of GFR was defined as a sustained decline in eGFR of more than 5 ml/min/1.73 m^2^/year.
- Rate of hospitalization, mortality, and total abdominal paracentesis times within 12 weeks were followed.

### Statistical analysis

The main endpoint of the study selected to calculate the sample size was the comparison of urine TIMP2 and IGFBP7 between the 3 liters and 5 liters abdominal paracentesis groups. It was calculated that 30 patients per group were needed to detect the difference of 20% in the proportion of patients showing the predictive performance of urine TIMP2 and IGFBP7 between the two groups with an α error of 5% and a β error of 10%.

Baseline clinical and biochemical data were compared using an independent t test or chi-square test, while categorical variables were compared using a repeated ANOVA test. Continuous variables were presented as mean ± standard deviation (SD) for normal distribution data, median with interquartile range (IQR) for non-normal distribution, while categorical variables were presented as percentage and frequency over the total available. The rapid decline of GFR at 12 weeks of follow-up was compared between groups using the Fisher exact test. Significantly was established at a P value of < 0.05. The analysis was done using IBM SPSS Statistics for Windows, Version 22.0 (IBM Corp., Armonk, NY).

## Results

### Study population

We screened 90 outpatients with decompensated cirrhosis from December 2018 to December 2021. Twenty-six patients were excluded, leaving 64 patients who were enrolled and randomized. Figure 1 summarizes the flowchart of the study. Patients who deviated from the intended liters of ascites release were also excluded. Leaving 29 patients in the 3-liter group and 25 patients in the 5-liter group, they were analyzed.

The baseline demographic characteristics of the study population were similar between the two groups, as shown in Table 1. The mean age of patients was 64 years, and most of the patients were male (87%). The most common cause of decompensated cirrhosis in the study was alcoholic cirrhosis (50%). Most patients had CTP class B cirrhosis (78%). The means of MELD and MELD-Na scores were 12.9±4.8 and 16.2±5.4, respectively. The median dose of spironolactone was 50 mg in both groups. There were no statistically significant differences in the underlying disease of patients or biochemical values between the two groups (Table 1).

**Table 1.** Demographic data*.

| **Variables** | **3 L group (n=29)** | **5 L group (n=25)** | ***p*-value** |
| --- | --- | --- | --- |
| **Age (Years)** | 64 ± 10.2 | 65 ± 10.96 | 0.75 |
| **Male** | 24 (82%) | 23 (92%) | 0.47 |
| **Body weight (kg)** | 63 ± 13.2 | 62. ± 15.2 | 0.86 |
| **BMI (Kg/m<sup>2</sup>)** | 23.3 ± 4.6 | 22.4 ± 5.3 | 0.51 |
| **Etiology** |  |  |  |
| **Alcohol** | 14 (48%) | 13 (52%) | 0.78 |
| **Hepatitis B virus** | 5 (18%) | 5 (20%) | 0.95 |
| **Hepatitis C virus** | 4 (14%) | 2 (8%) | 0.49 |
| **Nonalcoholic steatohepatitis** | 4 (14%) | 3 (12%) | 0.84 |
| **Others** | 2 (6%) | 2 (8%) | 0.78 |
| **Child-Pugh score** | 8.9 ± 1.4 | 8.8 ± 1.05 | 0.96 |
| **CTP B** | 22 (76%) | 20 (80%) | 0.71 |
| **CTP C** | 7 (24%) | 5 (20%) |  |
| **MELD score** | 12.2 ± 4.4 | 13.5 ± 5.1 | 0.33 |
| **MELD-Na score** | 15.1 ± 5.5 | 17.4 ± 5.2 | 0.13 |
| **Diuretic dosage** |  |  |  |
| **Spironolactone (mg)** | 50 (25, 50) | 50 (25, 100) | 0.22 |
| **Furosemide (mg)** | 20 (0, 20) | 20 (0, 40) | 0.40 |
| **Underlying disease** |  |  |  |

| Variables | 3 L group (n=29) | 5 L group (n=25) | *p*-value |
| --- | --- | --- | --- |
| **Diabetes** | 15 (51%) | 15 (60%) | 0.54 |
| **Hypertension** | 9 (31%) | 13 (52%) | 0.11 |
| **Dyslipidemia** | 4 (13%) | 6 (24%) | 0.33 |
| **Coronary artery disease** | 1 (3%) | 2 (8%) | 0.46 |
| **Biochemical value** |  |  |  |
| **Hemoglobin (g/dl)** | 10.2 ± 1.9 | 10.03 ± 1.5 | 0.68 |
| **Platelet (x10<sup>9</sup>/L)** | 189 ± 78 | 213 ± 83 | 0.27 |
| **INR** | 1.22 ± 0.2 | 1.25 ± 0.2 | 0.72 |
| **Albumin (g/dL)** | 2.8 ± 0.6 | 2.8 ± 0.4 | 0.77 |
| **Bilirubin (mg/dL)** | 1.6 ± 2.04 | 2.3 ± 4.1 | 0.40 |
| **Sodium (mEq/L)** | 134 ± 4 | 133 ± 4 | 0.44 |
| **Creatinine (mg/dL)** | 1.2 ± 0.4 | 1.4 ± 0.7 | 0.20 |
| **CKD-EPI (ml/min/1.73m<sup>2</sup>)** | 65 ± 28 | 62 ± 31 | 0.67 |

\*Value presented as mean ± SD. Or median (IQR) and n (%).
*P*-value corresponds to Independent t test and chi-square test.

### Primary outcome

The AKI event was assessed within 2 weeks after abdominal paracentesis with no statistical difference between the 3L and 5L groups, n = 3 (10%) vs. N = 3 (12%); *p*=0.85. (Table 2). All urinary biomarkers showed no statistical difference between AKI events and no AKI event (Table 3).

**Table 2.** Urine biomarkers and outcomes between two groups*.

| Variables | 3 L group (n=29) | 5 L group (n=25) | *p*-value |
| --- | --- | --- | --- |
| **TIMP2.IGFBP7/1000 >2\*\*** | 5 (17%) | 12 (48%) | 0.01* |
| **Rising Urine TIMP2** | 3 (10%) | 8 (32%) | 0.049* |
| **Rising Urine IGFBP7** | 8 (27%) | 8 (32%) | 0.72 |
| **Rising Urine TIMP2.IGFBP7** | 5 (17%) | 7 (28%) | 0.34 |
| **Rising Urine TIMP2/Urine Cr** | 12 (41%) | 19 (76%) | 0.01* |
| **Rising Urine IGFBP7/Urine Cr** | 12 (41%) | 13 (52%) | 0.43 |
| **Rising Urine TIMP2.IGFBP7/**<br>**Urine Cr** | 9 (31%) | 12 (48%) | 0.20 |
| **Acute kidney injury** | 3 (10%) | 3 (12%) | 0.85 |
| **Hemodynamic event** | 3 (10%) | 5 (20%) | 0.31 |
| **Change MAP (mmHg))** | -3.6±3.7 | -5.2±4.3 | 0.06 |
| **Ascites release times (times)** | 4 (3, 6) | 6 (4, 6) | 0.032* |
| **Rapid decline of GFR in 3 months** | 16 (55%) | 14 (56%) | 0.95 |
| **Admission within 3 months** | 7 (24%) | 8 (32%) | 0.52 |
| **Death within 3 months** | 3 (10%) | 2 (8%) | 0.76 |

\*Value presented as n (%). P-value corresponds to independent t test or Pearson Chi-Square test.
\*\*TIMP2 x IGFBP7)/1000 > 2 means that the result is positive, and patients are at risk of renal injury.

**Table 3.**
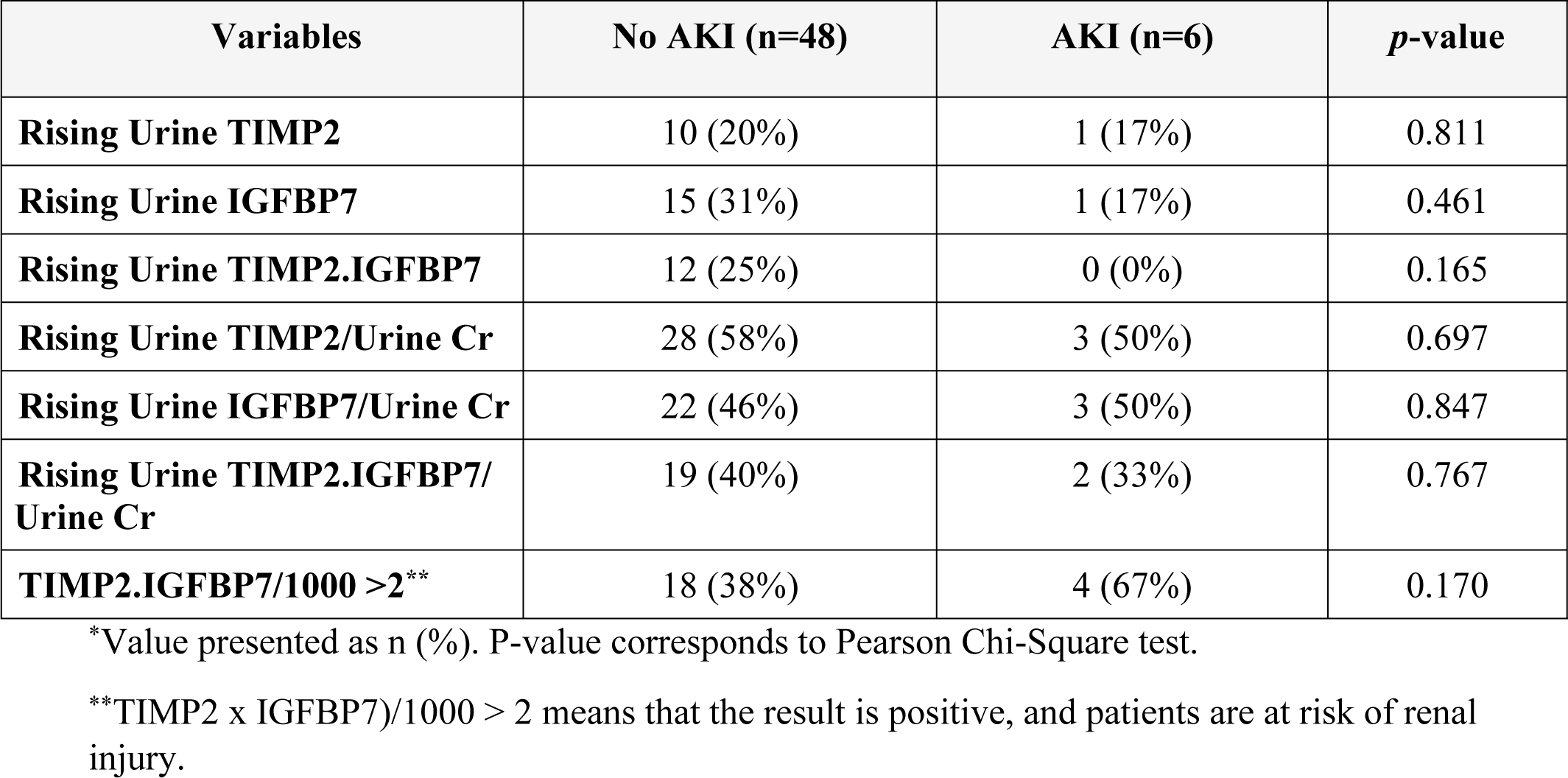
Prediction of AKI event by urinary biomarker*.

Statistically, rising urine TIMP2, rising urine TIMP2/urine Cr were all higher with a 5 L paracentesis (n = 5 (17%) vs. N = 12 (48%); p=0.01; n = 3 (10%) vs. N = 8 (32%); p=0.049; and n = 12 (41%) vs. N = 19 (76%); p=0.01, respectively. However, there was no statistically significant difference between the 3 liters and 5 liters groups in rising urine IGFBP7, urine TIMP2.IGFBP7, urine IGFBP7/urine Cr and urine TIMP2.IGFBP7/urine Cr. (Table 2).

### Secondary outcome

Hemodynamic event was assessed during abdominal paracentesis with no statistical difference between the 3 liters and 5 liters groups, n = 3 (10%) vs. N = 5 (20%); *p* = 0.31. The rapid decline of GFR events was not statistically different between the two groups, n = 16 (55%) vs. n = 14 (56%); *p* = 0.95. Ascites release times were statistically different between the 3 liters group = 4 (3, 6) and the 5 liters group = 6 (4, 6); *p* = 0.032. There was no statistical difference between 2 groups in admission and death within 3 months of the follow up (Table 2). Five patients died within 90 days of follow-up according to sepsis (n = 2), spontaneous bacterial peritonitis (n = 2) and primary bacteremia (n = 1).

Rising urine IGFBP7, rising urine TIMP2.IGFBP7, rising urine IGFBP7/urine Cr, rising urine TIMP2.IGFBP7/ urine Cr and urine TIMP2xIGFBP7/1000 > 2 were statistically significantly higher hemodynamic events than without increases hemodynamic events, n = 9 (19%) vs. n = 7 (78%); *p>*0.001, n = 6 (13%) vs. n = 6 (75%); *p>*0.001, n = 18 (39%) vs. n = 7 (87%); p=0.011, n = 14 (30%) vs. n = 7 (87%); p=0.002 and n = 11 (23%) vs. n = 6 (75%); *p*=0.004, respectively (Table 4).

**Table 4.**
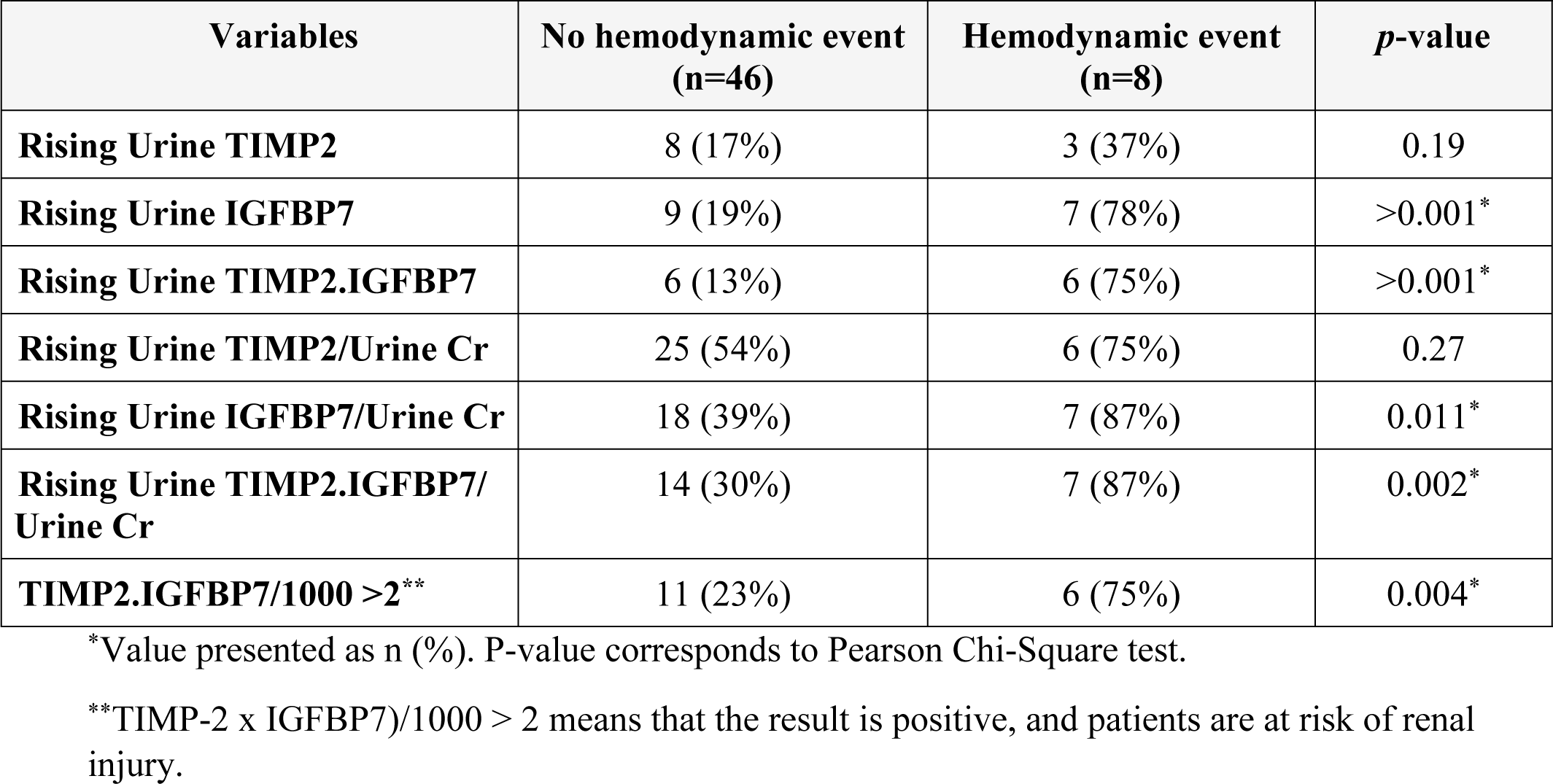
Prediction of hemodynamic event by urinary biomarker*.

Mean arterial pressure was statistically significantly declined at 2 hours of abdominal paracentesis compared with baseline within both groups, 3 liters group declined 3.6 (95% CI: 1.8-5.4), P<0.001 and 5L group declined 5.2 (95% CI: 2.9-7.6), P<0.001. Between the 2 groups there was no statistically significant decline. (Figure 2) Urine TIMP2.IGFBP7 more than 3.435 (AUC 0.85, 95% CI 72-0.99; *p* = 0.002), rising urine TIMP2.IGFBP7/ urine Cr more than 0.0027 (AUC 0.84, 95% CI 0.65-1; *p* = 0.002) and rising urine IGFBP7/urine Cr more than 0.0025 (AUC 0.80, 95% CI 0.06-0.95; *p* = 0.006) were statistically significant to predict a hemodynamic event. (Figure 3).

**Figure 2.**
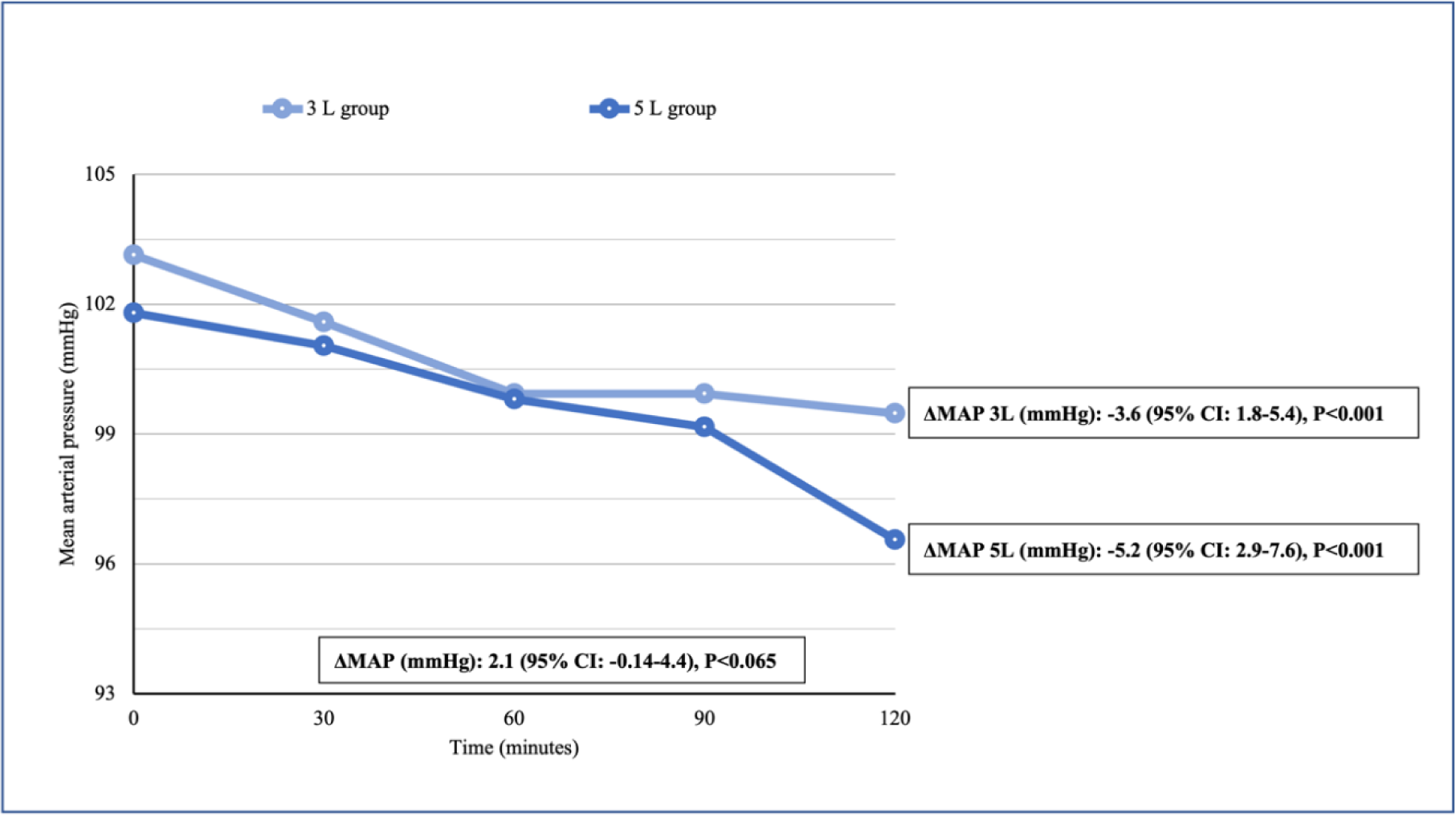
Declined mean arterial blood pressure between two groups Comparing mean arterial pressure between groups. \**P*-value was less than 0.05.

**Figure 3.**
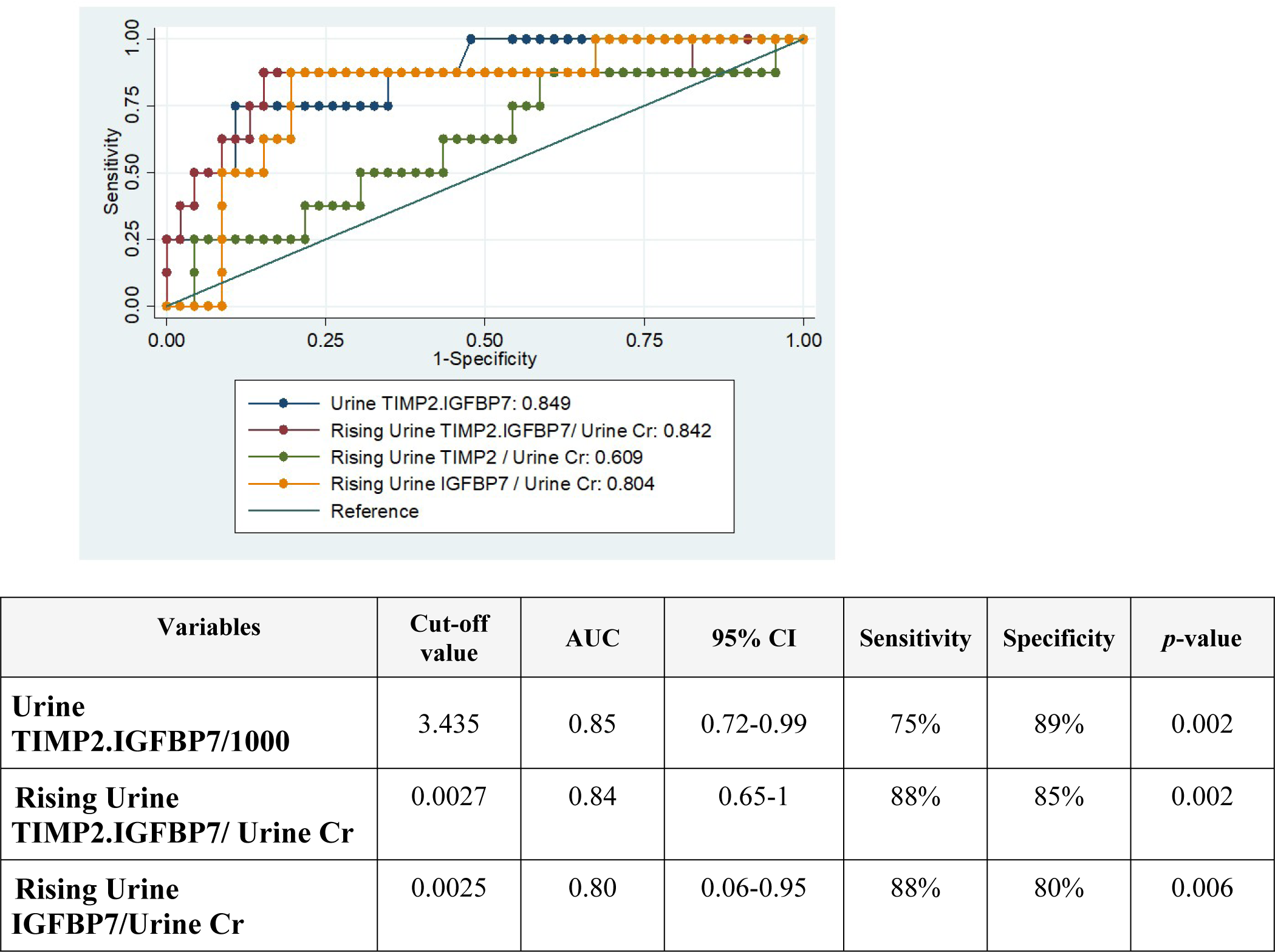
ROC curve for predicting Hemodynamic event.

## Discussions

This study has four key findings. First, urine TIMP2.IGFBP7/1000 >2 shows the good performance in predicting the rapid decline of GFR and hemodynamic events. Second, urinary TIMP2 and IGFBP7 can be more sensitive for early detection of tubular injury compared with serum creatinine. Third, the rapid decline of GFR in 90 days is high in both the 3 and 5 L groups of abdominal paracentesis. Fourth, renal tubular injury can occur even when less than 5 liters of ascites are released in decompensated cirrhosis.

Evaluation of acute kidney injury by using serum creatinine has several limitations. Novel biomarkers have displayed their role for early detection in AKI in many recent studies. Urine TIMP2.IGFBP7/1000 >2 in several studies has demonstrated the usefulness of this biomarkers in predicting AKI^14–16^, but there have been no studies in cirrhotic patients. The hypothesis of this study is presented in figure 4.

**Figure 4.**
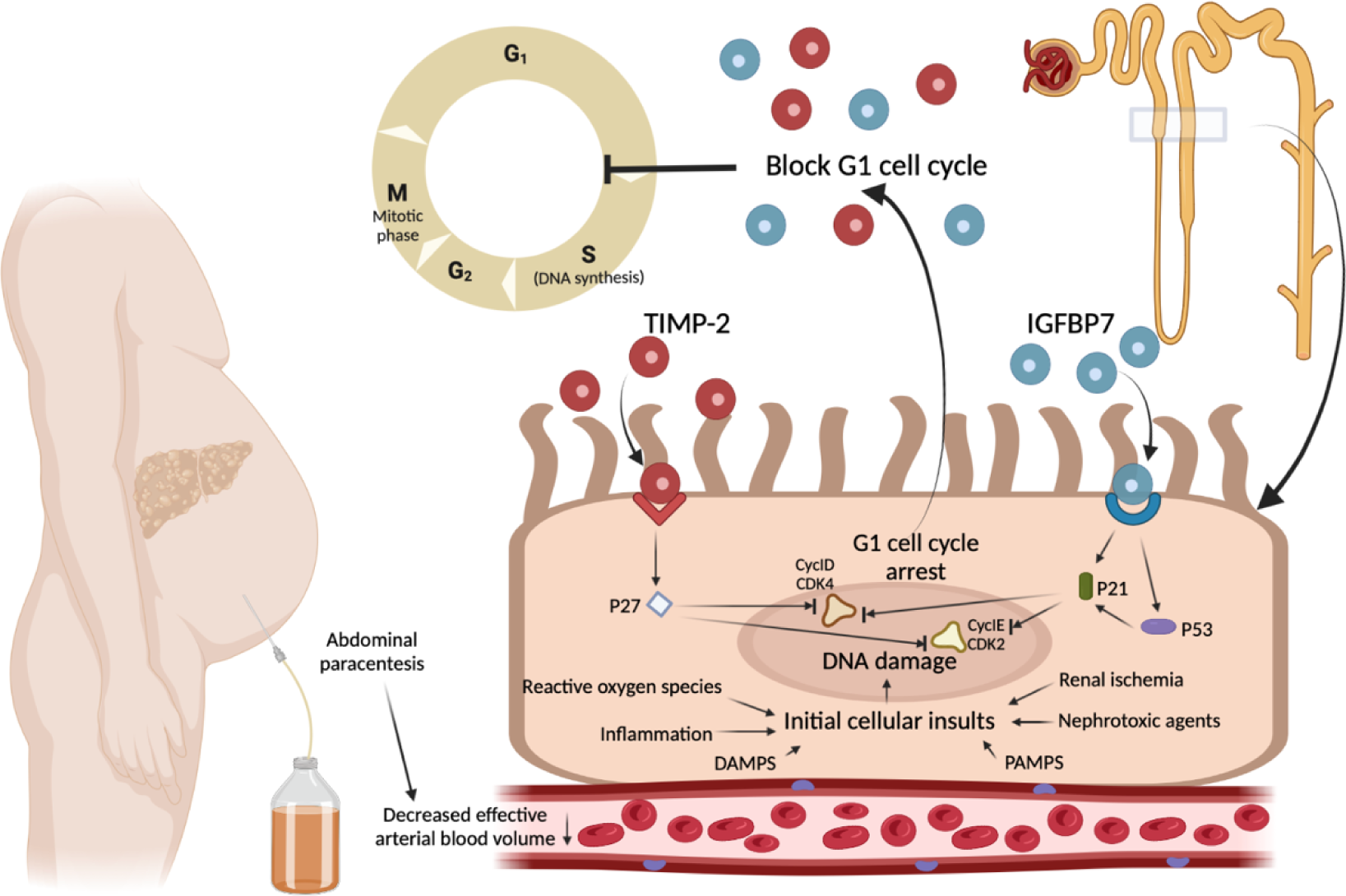
The hypothesis of renal tubular injury with detection of TIMP2 and IGFBP7 in cirrhosis. Adapted from Kashani K, *et al*.14 Proposed mechanism of the novel biomarkers in AKI developing in patients with cirrhosis: Initial renal tubular injury by decreased effective arterial blood volume during abdominal paracentesis. Regarding to DNA of tubular cellular damage, IGFBP7 and TIMP2 are secreted in the tubular cells. IGFBP7 directly increases the expression of p53 and p21 and TIMP-2 stimulates p27 expression via IGFBP7 and TIMP2 receptors. Theses p proteins then block the effect of the cyclin-dependent protein kinase complexes (CyclD-CDK4 and CyclE-CDK2) on the cell cycle promotion, thereby resulting in G_1_ cell cycle arrest for short periods of time presumably to avoid cells with possible damage from dividing.

Adapted from Kashani K, *et al*.^14^ Proposed mechanism of the novel biomarkers in AKI developing in patients with cirrhosis: Initial renal tubular injury by decreased effective arterial blood volume during abdominal paracentesis. Regarding to DNA of tubular cellular damage, IGFBP7 and TIMP2 are secreted in the tubular cells. IGFBP7 directly increases the expression of p53 and p21 and TIMP-2 stimulates p27 expression via IGFBP7 and TIMP2 receptors. Theses p proteins then block the effect of the cyclin-dependent protein kinase complexes (CyclD-CDK4 and CyclE-CDK2) on the cell cycle promotion, thereby resulting in G_1_ cell cycle arrest for short periods of time presumably to avoid cells with possible damage from dividing.

This is the first randomized prospective single center study that has explored the value of both novel biomarkers, TIMP2 and IGFBP7 for predicting rapid decline of GFR, hemodynamic event, 90-day admission, and 90-day mortality in decompensated liver cirrhotic patients who had been diagnosed with diuretic-resistance or diuretic-intractable ascites and randomly received moderate volume paracentesis for 3 liters and 5 liters groups. For patients presenting with tense ascites, large-volume paracentesis (LVP) combined with hyper oncotic human albumin is the initial treatment of choice. In patients with diuretic-resistance or diuretic-intractable ascites, albumin infusion may be considered for paracentesis of a smaller volume to prevent further complications.

In this study, the etiology of cirrhosis was the majority of alcohol (50%), which was comprised of chronic HCV (11%), chronic HBV (20%), NASH (13%) and autoimmune (3.5%). The mean CTP score was 8.9 which is similar to the study of Belcher JM, et al^9^, which aimed to evaluate structural (neutrophil gelatinase–associated lipocalin, IL-18, kidney injury molecule-1 [KIM-1], liver-type fatty acid–binding protein [L-FABP], and albuminuria) and functional (fractional excretion of sodium [FENa]) urinary biomarkers as predictors of AKI progression and in-hospital mortality.

The median dose of spironolactone and furosemide was 50 mg and 20 mg, respectively. The dose of spironolactone and furosemide was not at a maximum dose due to the fact that most decompensated cirrhotic patients in this study had diuretic intractable problems. At this point, the dosage of diuretics could not affect the kinetics of kidney function.

Although 16 (55%) and 14 (56%) of the 54 decompensated cirrhotic patients received 3 liters and 5 liters of abdominal paracentesis, respectively, they developed rapid declines in GFR in 90 days, and creatinine with a lack of sensitivity to this parameter of renal damage in cirrhotic patients. In contrast, urine TIMP2.IGFBP7/1000 >2 and rising TIMP2 were significantly found in the 5 L group. This is comparable to the results of Bihorac A, et al ^18^ which demonstrated urine TIMP2.IGFBP7/1000 >2 predicted AKI in ICU patients who received major operations and nephrotoxic agents.

The performance of urine TIMP2.IGFBP7 as a predictor of the rapid decline of GFR events was superior to that of serum creatinine. This finding suggests that this renal tubular injury marker may be more sensitive for early detection of the rapid decline of renal function in advanced cirrhosis.

Urine TIMP2.IGFBP7/1000, rising urine TIMP2.IGFBP7/Cr and rising urine IGFBP7/urine Cr could predict hemodynamic events with AUC of 0.85 (95% CI 72-0.99), 0.84 (95% CI 0.65-1) and 0.80 (95% CI 0.06-0.95), respectively. The predictive performance of TIMP2.IGFBP7 was as good as the other biomarkers such as urine NGAL (AUC 0.77; 95% CI: 0.68-0.85), urine IL-18 (AUC 0.71; 95% CI 0.61-0.81), urine KIM-1 (AUC 0.66; 95% CI 0.56-0.76) and urine L-FABP (AUC 0.76; 95% CI; 0.66-0.85) as reported in the study of Belcher JM, et al^9^.

Rising urine IGFBP7, rising urine TIMP2.IGFBP7, rising urine IGFBP7/urine Cr, rising urine TIMP2.IGFBP7/urine Cr and urine TIMP2.IGFBP7/1000 > 2 were statistically significant for higher hemodynamic events than without increases in hemodynamic events in both the 3-liter and 5-liter groups. It is confirmed that renal tubular injury will increase in people with abdominal paracentesis of less than 5 liters. This is the strength of this study that defines the performance of two novel biomarkers, which the previous two studies from Bihorac A, et al^15^ and Hoste EA, et al^16^ did not perform. This finding confirmed that TIMP2 and IGFBP7 are known to be involved in the response to a wide variety of insults such as inflammation, ischemia, oxidative stress, or receiving renal toxicity agents.^11–13^ This may help explain why they correspond to the risk of AKI in patients with decompensated cirrhosis. AKI is one of the clinical manifestations of post-paracentesis circulatory dysfunction (PPCD).

According to the EASL clinical practice guidelines for the management of patients with decompensated cirrhosis, in patients undergoing paracentesis of less than 5 liters of ascites, the risk of developing PPCD is low. Our study revealed renal tubular injury, even though less than 5 liters of ascites were removed. Our finding might support the use of albumin infusion in patients receiving paracentesis of less than 5 liters of ascites to prevent expanding PPCD.

Most patients in this study demonstrated hemodynamic events (15%) and rapid decline of GFR events (55%), and they were not statistically different between the two groups. This data has shown that even with an abdominal paracentesis of less than 5 liters, a decline in mean arterial pressure and further damaged to the kidney can occur. This is in contrast to the previous study by Peltekian KM, et al^17^ which demonstrated a single 5 liters paracentesis in patients with cirrhosis and tense, diuretic-resistant ascites without albumin infusion caused no disturbances in systemic and renal hemodynamics 48 hours after the procedure. These findings could support the conclusion that although short-term hemodynamic and renal outcomes were not achieved, the rapid decline of GFR in 90 days can be displayed even if an abdominal paracentesis of less than 5 liters was performed.

There were some limitations to this study. First, this is a single-center study with a relatively small sample size, and only enrolled patients with diuretic-resistance or diuretic-intractable ascites. Further studies are recommended to confirm this result and its application in clinical practice. Second, this study recruited only outpatient decompensated cirrhosis patients, so these results cannot be applied to all cirrhotic patients, including acute decompensated cirrhosis and acute-on-chronic liver failure. Third, the urine biomarker data was not performed in cirrhotic patients who received more than 5 liters of ascites release. Finally, this study did not validate the model to predict the risk of a rapid decline in GFR, hemodynamic event or 90-day mortality in an external cohort. However, this result is the first report that has confirmed the effectiveness of urine TIMP2 and IGFBP7 and also confirmed that those who received moderate volume paracentesis actually caused renal tubular injury. Future research may quantify the key hormone that plays a role in the pathophysiology of the accumulation of ascites by measuring plasma levels of renin and aldosterone that may be collinear with the volume of ascites release and a rise in urinary TIMP2 and IGFBP7.

### Conclusions

Urine TIMP2.IGFBP7>2 predicted renal tubular injury in patients who had moderate volume paracentesis, which mean that releasing ascites below 5 liters may not be safe. Kidney injury could occur when even less than 5 liters of ascites release were performed in decompensated cirrhotic patients.

## Clinical trial registration

This study was approved by the institutional review board and was registered with the national clinical registry; the registration number was **TCTR20191116003**. The full trial protocol can be accessed at www.clinicaltrials.in.th.

## Data Availability

The data that support the findings of this study are available on request from the corresponding author, [SC]. The data are not publicly available due to their containing information that could compromise the privacy of research participants.

## Acknowledgement

The authors wish to thank Mr. Stephen Pinder, a native-speaking medical English specialist from the Department of Clinical Epidemiology and Biostatistics, Faculty of Medicine, Ramathibodi Hospital, Mahidol University for English review and editing.

## Ethics approval and consent to participate

All subjects were properly instructed and consented to participate in this trial by signing the informed consent regulation provided by the Institutional Review Board of the Royal Thai Army Medical Department Committee (IRB number R164h/61) and registered at www.clinicaltrials.in.th (TCTR20191116003). The Institutional Review Board of the Royal Thai Army Medical Department Committee uses the World Medical Association: DELCARATION OF HELSINKI, GUIDELINES FOR GOOD CLINICAL PRACTICE: ICH Harmonized Tripartite Guideline, Council for International Organizations of Medical Sciences (CIOMS), CODE of FEDERAL REGULATIONS: Title 45 Public Welfare; Part 46 Protection of Human Subjects and the Belmont Report to regulate the ethics concerns in publications.

Informed consent was obtained from all subjects, and all methods were conducted according to the relevant guidelines and regulations.

## Consent for publication

Informed consent was obtained by signature of all participants and from all subjects to provide all the information regarding publication.

## Availability of data and materials

The datasets used during the current study are available from the corresponding author on reasonable request.

## Authors’ contributions

Drs. Chirapongsathorn and Suksamai had full access to all the data in the study and take responsibility for the integrity of said data and the accuracy of the data analysis.

Dr. Chirapongsathorn, Suksamai reviewed data for eligibility, evaluation of quality and data extraction, writing this manuscript, data analysis, and authorship.

Dr. Chirapongsathorn was supervisor and conceived the study concept.

All authors reviewed the data for eligibility and data extraction, evaluation of quality.

